# A novel DNA methylation-based surrogate biomarker for chronic systemic inflammation (InfLaMeS): results from the Health and Retirement Study

**DOI:** 10.1101/2024.10.11.24315339

**Authors:** Helen C.S. Meier, Eric T. Klopack, Mateo P. Farnia, Belinda Hernandez, Colter Mitchell, Jessica D. Faul, Cathal McCrory, Rose Anne Kenny, Eileen M. Crimmins

## Abstract

Chronic low-grade systemic inflammation is a risk factor for chronic diseases and mortality and is an important biomarker in health research. DNA methylation (DNAm) surrogate biomarkers are valuable exposure, risk factor and health outcome predictors in studies where the measures cannot be measured directly and often perform as well or better than direct measure. We generated a DNAm surrogate biomarker for chronic, systemic inflammation from a systemic inflammation latent variable of seven inflammatory markers and evaluated its performance relative to measured inflammatory biomarkers in predicting several age-associated outcomes of interest, including mortality, activities of daily living and multimorbidity in the Health and Retirement Study (HRS). The DNAm surrogate, Inflammation Latent Variable Methylation Surrogate (InfLaMeS), correlated with seven individual inflammation markers (r= −0.2-0.6) and performed as well or better to the systemic inflammation latent variable measure when predicting multimorbidity, disability, and 4-year mortality in HRS. Findings were validated in an external cohort, The Irish Longitudinal Study of Ageing. These results suggest that InfLaMeS provides a robust alternative to measured blood-chemistry measures of inflammation with broad applicability in instances where values of inflammatory markers are not measured but DNAm data is available.

## INTRODUCTION

Inflammation has been identified as a key biomarker to investigate health and health inequalities within individuals and across populations. Inflammatory indicators have consistently been shown to be socially patterned, with those of lower socioeconomic status having higher levels of inflammation.^1–3^ Chronic inflammation has also been tied to several age-related conditions and diseases, including cardiovascular disease^4^, diabetes^5,6^, stroke^7^, cognitive decline^8^, frailty^9^ and mortality^10-12^. Most existing studies have heavily relied on levels of various blood-based markers of inflammation including C-Reactive Protein (CRP) and individual cytokines such as Interleukin-6 (IL-6. Specifically, elevated levels of CRP and IL-6 have consistently been associated with greater all-cause mortality as well as cardiovascular and cancer-related mortality^11,13^ and IL-6 has been demonstrated to be a marker of frailty in cohort studies.^14^

Prior studies of inflammation among older adult populations have largely relied on individual measures (or composites of a few measures) of inflammatory markers. While these approaches have provided advancements in understanding how inflammation may be a result of social exposures and related to later life chronic conditions and diseases, they are limited in that individual markers may be tied to specific outcomes and capture specific inflammatory processes that may lead to mixed findings and does not fully capture complex cascades, feedback loops, nor the extent of inflammation risk within individuals that may be system wide. Efforts to generate composite measures of systemic inflammation have struggled with how to harmonize different sets of inflammatory markers and vary by the availability of measured biomarkers and the manner in which these indices are aggregated to create biological risk profiles. For example, common methods include characterizing risk by creating decile groups for each inflammatory biomarker and then sum across deciles to create a risk profile^15^ and summing z-score values of inflammatory markers^16^. Clinically focused scores, such as the systemic inflammation grade^17^ and the Glasgow Prognostic Score^18^ categorize clinically relevant levels for each individual marker and then combine levels of risk. While these composites do predict important health outcomes, including cancer, depression and mortality, the variation in composite score generation and biomarkers contributing to the scores limits reproducibility and doesn’t account for covariation of biomarkers. Additionally, many studies do not measure inflammatory markers and are therefore unable to assess this critical aging and health pathway.

Recently, a growing body of research has turned to markers based on DNA methylation (DNAm) to predict age-related outcomes.^19^ DNA methylation, cytosine-5 methylation at CpG sites, is an important epigenetic mechanism contributing to gene expression, and changes in DNAm that occur with age are considered one of the hallmarks of aging.^20^ DNAm composite scores are derived from CpG sites that predict age and age-related phenotypes, the most famous of which are epigenetic clocks.^19^ Epigenetic clocks, surrogates of age or age-related phenotypes, have become popular in studies of environmental, social and biological determinants of risk years before onset of health outcomes.^21^ A parallel application of DNAm scores is to generate surrogates for exposures, risk factors^22,23^ and specific health outcomes.^24^ DNAm surrogates can be calculated in any study with existing DNAm data, facilitating replication of results, meta-analyses, and the use of DNAm surrogates to generate proxies of factors that were not originally measured. Further, DNAm surrogates can perform as well or better than measured exposures when predicting outcomes.^9,25,26^

Previous studies have generated DNAm surrogates for levels of individual biomarkers, such as CRP^27^ and IL–6.^28^ The goal of this study was to generate a DNAm surrogate biomarker for chronic systemic inflammation, using a previously generated latent variable that leveraged seven inflammatory indicators to more holistically capture systemic inflammation.^10^ We then evaluated the performance of the systemic Inflammation Latent Variable Methylation Surrogate (InfLaMeS) relative to measured biomarkers for chronic systemic inflammation in predicting several age-associated outcomes of interest, including mortality, activities of daily living and multimorbidity in two independent cohort studies of older adults.

## METHODS

Analyses were conducted in three phases (Figure 1): 1) training (development of the surrogate), 2) testing (examination of the surrogate within a hold-out sample), and 3) replication in an external study.

**Figure 1.**
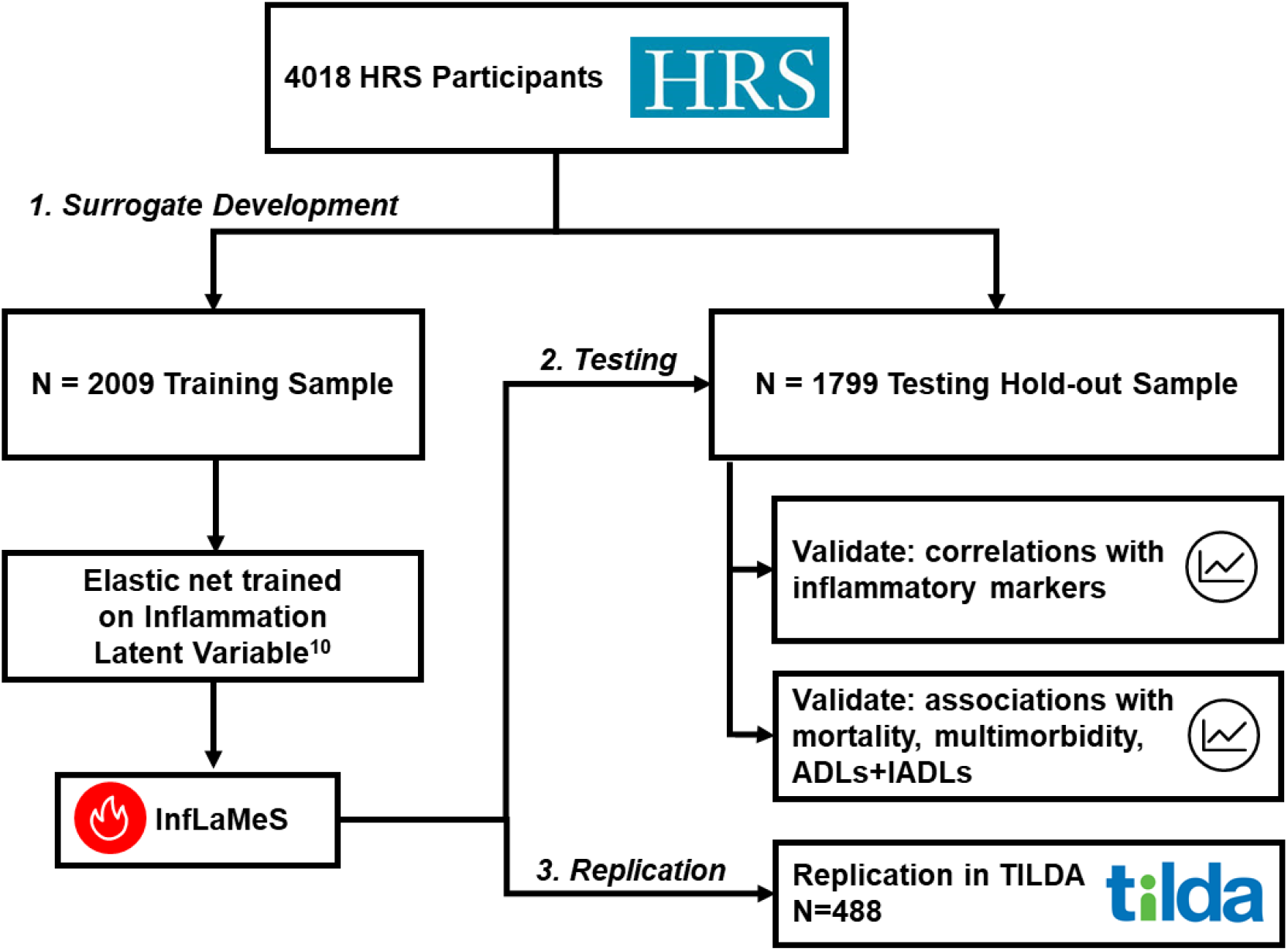
Plan of analysis.

### Training and testing study population

The Health and Retirement Study (HRS) is a nationally representative sample of US adults over the age of 50 years. The 2016 Venous Blood Study (VBS) is a subsample of 9,934 HRS participants who completed an interview during the 2016 HRS wave and had venous blood collected by a trained phlebotomist at a subsequent in-home visit.^29^ As described previously, systemic inflammation latent variable was generated from 9,873 participants who had at least one of seven biomarkers of inflammation.^10^ DNA methylation (DNAm) was measured on a subsample of 4,104 VBS participants. Eighty-six individuals were excluded for failing quality control procedures, resulting in a sample of 4,018 for the DNAm surrogate creation. Missing CpG sites were median imputed and then this sample was randomly split into a training set (N=2,009) and a testing set (N=2,009) to evaluate the surrogate performance predicting age-associated health outcomes.

### Systemic inflammation biomarker

Following Meier et al., the latent variable representing overall systemic inflammation was created from the 6 log-transformed cytokine measurements, C-reactive protein (CRP, referent), Interleukin-6 (IL-6), Interleukin-10 (IL-10), Interleukin-1 receptor antagonist (IL-1ra), Insulin-Like Growth Factor 1 (IGF-1), and Tumor Necrosis Factor-1 (sTNFR-1), and the neutrophil to lymphocyte ratio.^10^ Inflammatory markers assayed from VBS participant serum at the University of Minnesota Advanced Research and Diagnostic Laboratory as described previously.^29^ Neutrophil-to-lymphocyte ratio was derived from flow cytometry data by dividing the percent of neutrophils by the percent of lymphocytes.

### DNA methylation measurement

DNA methylation was measured from assays using the Infinium Methylation EPIC BeadChip. Assays were completed at the University of Minnesota as described previously.^30^ Samples were randomized across plates by key demographic variables (i.e., age, cohort, sex, education, and race/ethnicity) with 39 pairs of blinded duplicates. Analysis of duplicate samples showed a correlation > 0.97 for all CpG sites. The *Minfi* package in R software, a suite of computational tools used to support preprocessing and quality control of Infinium Methylation EPIC BeadChip DNAm data^31^, was used for HRS DNAm data preprocessing and quality control. Sex mismatched samples and any controls (cell lines, blinded duplicates) were dropped. *Minfi* flagged 3.4% of the methylation assay probes (*n* = 29,431 out of 866,091) for suboptimal performance (i.e., methylated + unmethylated DNA signal at a given position not different than the background signal level from a negative control) using a detection *P*-value threshold of 0.01. *Minfi* flagged 58 samples using a 5% cutoff for detection *P*-value failed samples, and these samples were removed from the final dataset. High-quality methylation data is available for 97.9% of the samples (*n* = 4,018). DNAm information was provided for 833,865 CpG sites.

### Training the DNA methylation surrogate for systemic inflammation

We used an elastic net to create the systemic Inflammation Latent Variable Methylation Surrogate (InfLaMeS) in R 4.4.0 “Puppy Cup”^32^ using the glmnet package^33^. First, we divided the HRS VBS epigenetics subsample into training and testing sets of 2,009 participants each. Next, with the training set, we used an elastic net to determine which combination of CpG sites provided the best prediction of systemic inflammation. We selected the alpha and lambda hyperparameters for the elastic net using a grid search approach. 0.00, 0.10, 0.50, 0.70, 0.90, 0.95, 1.00 were included as possible alpha values. In this case, we found that an alpha of 0.1 and lambda of 0.184 provided a model with the lowest mean squared error. This model selected (i.e., produced non-zero coefficients for) 791 probes. We then estimated InfLaMeS in the testing set using the intercept, probes and coefficients produced by the model selected in the training set. InfLaMeS values ranged from [-0.75-1.23]. The coefficient values for CpG sites and code needed to produce InfLaMeS are available at the following GitHub (blinded).

### Health outcomes

We selected four age-associated health outcomes to evaluate the performance of InfLaMeS: 4-year mortality, activities of daily living (ADLs), instrumental activities of daily living (IADLs) and multimorbidity. Four-year mortality represented whether a participant was known to be deceased by HRS in 2020. The count of some difficulty with ADLs and IADLs reported at the 2016 interview were summed. The sum of positive responses from the 2016 interview to whether a doctor has ever told the respondent that they have ever had a condition for eight morbidities, including high blood pressure, diabetes, cancer, lung disease, heart disease, stroke, psychiatric problems and arthritis, was generated. Multimorbidity was categorized as yes (2 or more conditions) or no (0 or 1 conditions).

### Covariates

Covariates included chronologic age, race/ethnicity, gender, smoking status, body mass index (BMI) and heavy drinking. Age was measured in years. Race/ethnicity was categorized as non-Hispanic White, non-Hispanic Black, Hispanic and non-Hispanic Other and gender was dichotomized as male or female. Smoking status was classified as current, former or never smoker. Alcohol consumption was categorized into non-drinker, 1-4 drinks per day and 5+ drinks per day. The association of InfLaMeS with three epigenetic clocks was evaluated in this study, including GrimAge, PhenoAge and DunedinPACE. GrimAge and PhenoAge were adjusted for CpG-level principal components by the Higgins-Chen et al. method.^34^ There is no CpG-level principle components adjusted measure available for DunedinPACE.

### Testing analyses

In the hold-out testing sample, analyses were conducted to evaluate the performance of InfLaMeS. For these analyses, only participants who were not missing on covariates and had valid HRS survey weights were included (N = 1799). First, the correlations between InfLaMeS and the systemic inflammation latent variable, individual inflammatory markers (CRP, IL-10, IL-1RA, IL-6, TNFR-1, IGF-1 and neutrophil-lymphocyte ratio), principal components epigenetic clocks for GrimAge, PhenoAge, and DunedinPACE were calculated. Then, associations between InfLaMeS and age-associated health outcomes of interest (mortality, ADLs + IADLs, multimorbidity) adjusting for age, race/ethnicity, gender, smoking, BMI and alcohol consumption were examined. Logistic regression was used for 4-year mortality and cross-sectional multimorbidity; Poisson regression was used for the sum of ADLs and IADLs. The inflammation latent variable and InfLaMeS values were z-scored for comparability. Analyses were weighted using survey weights provided by HRS for the methylation subsample. In cases where a stratum has only one primary sampling unit (PSU; due to splitting data) the stratum contribution to the variance is the mean of all other strata with more than one PSU.^35^ All analyses were performed in R using the tidyverse^36^, jtools^37^ and survey^35^ packages.

Of note, while several other studies often control for cell type, we do not adjust for cell proportion in our study because cell type proportions are one of the indicators in the latent inflammation factor. Changes in cell type are part of the inflammation and inflammaging process are therefore part of the phenomena InfLaMeS is designed to capture. We therefore believe it is inappropriate to control for cell type.

### Replication

Coefficients for the 791 CpG sites included in the DNAm inflammation surrogate estimated in HRS data were applied to DNAm data (N=488) from The Irish Longitudinal Study on Ageing (TILDA). All inflammatory markers were log transformed and standardized. Correlations for the InfLaMeS and observed individual inflammation markers (IL-1RA, TNFR-II, CRP, IL-6, IGF-1 and IL-10) were estimated. Associations between InfLaMeS and mortality, ADLs/IADLs and multimorbidity adjusting for age, sex, smoking, obesity, and alcohol consumptions were replicated in TILDA using survey data. Logistic regression was used for 1) 12-year mortality, 2) presence of multimorbidity (two or more of the following conditions: heart disease, diabetes, hypertension, stroke or transient ischemic attack, lung disease, arthritis and cancer) and 3)

Presence of a disability (ADL or IADL). Poisson regression was further used to model the total number of ADLs/IADLs.

## RESULTS

Sample characteristics of the HRS testing and training data sets are shown in Table 1. The training data had a significantly lower proportion of those self-reporting as non-Hispanic other compared to those self-reporting as Black or non-Hispanic White, more current smokers, and fewer morbidity obese participants (*p* < .05 from a *t*-test). However, given the large number of *t*-tests and the randomization procedure it is highly likely these differences are due to chance.

**Table 1.** Weighted demographic characteristics (mean or percent) of the Health and Retirement Study training and testing samples.

| Sample characteristics of testing and training sets in HRS |  |  |
| --- | --- | --- |
|  | Testing Data | Training Data |
| Age (years) | 68.32 | 68.77 |
| Race/ethnicity |  |  |
| White, not Hispanic | 0.79 | 0.81 |
| Black, not Hispanic | 0.10 | 0.11 |
| Hispanic | 0.06 | 0.06 |
| Other race, not Hispanic | 0.05 | 0.02 |
| Female | 0.55 | 0.52 |
| Smoking Status |  |  |
| Current Smoker | 0.10 | 0.12 |
| Never Smoked | 0.46 | 0.43 |
| Past Smoker | 0.44 | 0.44 |
| Body Mass Index (BMI) |  |  |
| Under Weight | 0.02 | 0.02 |
| Normal Weight | 0.25 | 0.26 |
| Overweight | 0.35 | 0.38 |
| Obese | 0.23 | 0.22 |
| Morbidly Obese | 0.15 | 0.12 |
| Alcohol Use |  |  |
| Non-Drinker | 0.57 | 0.56 |
| 1-4 Drinks per day | 0.40 | 0.42 |
| 5+ Drinks per day | 0.03 | 0.03 |

We first examined the association between InfLaMeS, the inflammation latent variable and individual markers. The weighted correlation between InfLaMeS and the observed inflammation latent variable was 0.632 (Figure 2). A correlation matrix for the inflammation latent variable, InfLaMeS, individual inflammatory markers and epigenetic clocks is presented in Table 2. The DNAm inflammation surrogate was positively correlated with CRP (r=0.45), IL-10 (0.31), IL1-RA (0.43), IL-6 (0.43), TNFR-1 (0.49), and neutrophil-lymphocyte ratio (0.59), and negatively correlated with IGF-1 (−0.20). InfLaMeS was positively correlated with all three epigenetic clocks (PCGrimAge r =0.49; PCPhenoAge r=0.61; DunedinPACE r=0.62). Correlation estimates for the InfLaMeS and individual inflammatory markers were smaller than correlations between the inflammation latent variable and individual inflammatory markers, while correlations between epigenetic clocks and InfLaMeS and between InfLaMeS and neutrophil-lymphocyte ratio were stronger than for the inflammation latent variable. In the testing sample, controlling for age, race/ethnicity and gender, the InfLaMeS was associated with the observed inflammation latent variable (b = 1.18, *p* < 0.001). In bivariate regressions conducted in the testing sample, InfLaMeS was associated with age (b = 0.01, *p* < .001) and being a member of a non-Hispanic other race (b = −0.11, *p* < .01), but not with gender or being a member of any other race/ethnic group.

**Table 2.**
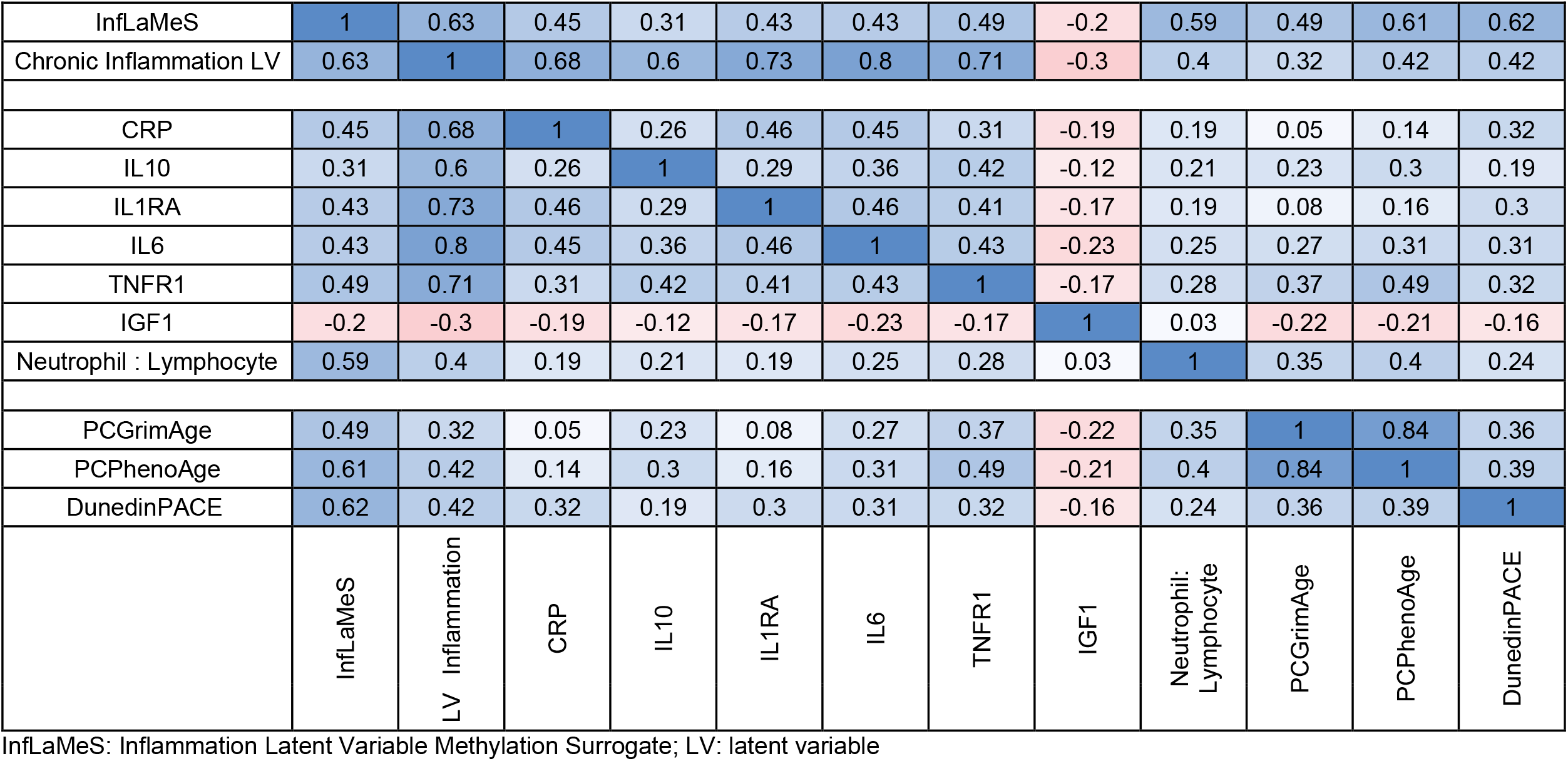
Correlation matrix of InfLaMeS, inflammation latent variable, individual biomarkers and three epigenetic clocks in the Health and Retirement Study testing sample (N=1799)

**Figure 2.**
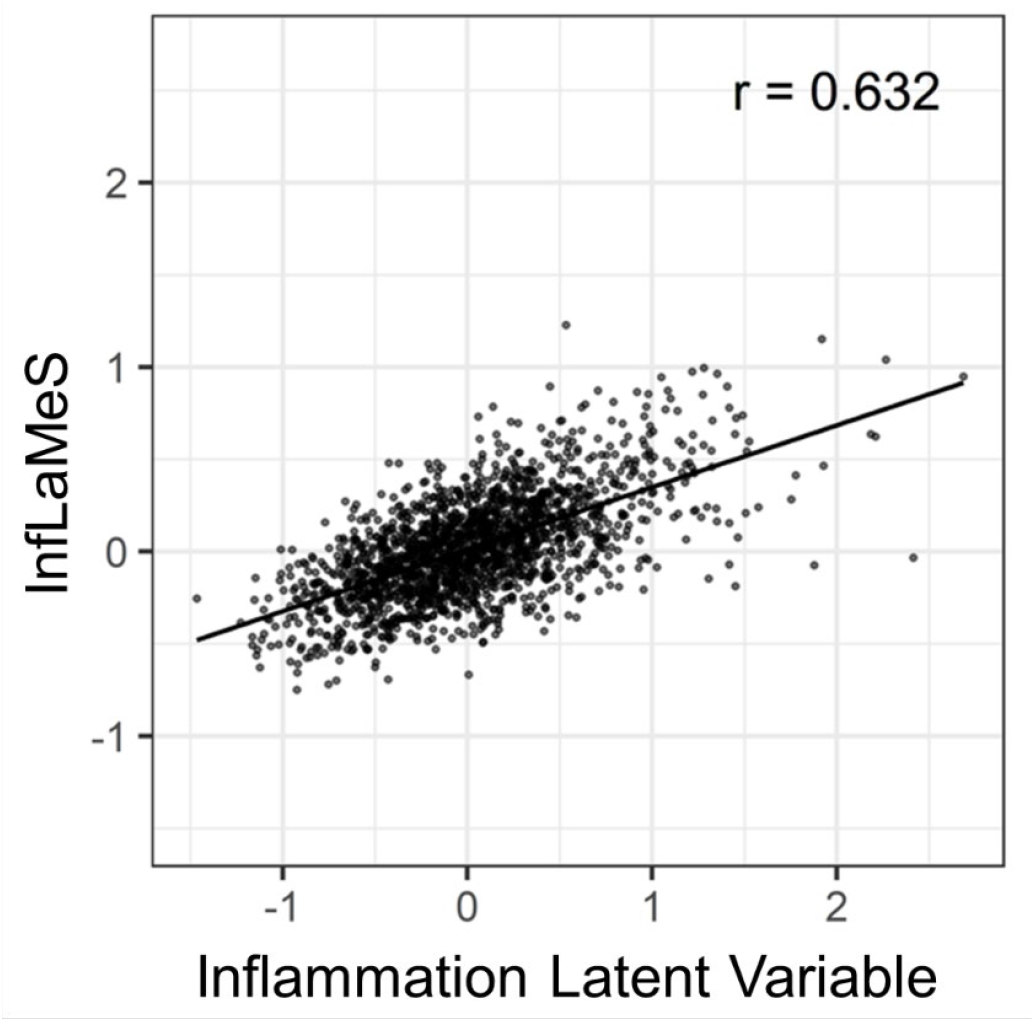
Weighted correlation between observed inflammation and DNAm surrogate in the Health and Retirement Study test sample (N= 1799).

Table 3 shows the weighted associations between 1) InfLaMes and 2) the inflammation latent variable and three health outcomes of interest: 4-year mortality, multimorbidity and the sum of activities of daily living and instrumental activities of daily living. For these models, both scores were standardized (z-scored) to facilitate comparison. All models were adjusted for age, gender, race/ethnicity, smoking status, obesity and alcohol consumption. The InfLaMeS had higher effect estimates for mortality (OR=2.24, 95%CI= 1.80, 2.73) and multimorbidity (OR= 1.75, 95% CI = 1.53, 2.02) than the measured latent variable (mortality OR=1.78, 95%CI=1.52, 2.08; multimorbidity OR=1.42, 95% CI=1.25, 1.53). The association between the inflammation latent variable and ADLs and IADLs was slightly stronger than InfLaMeS (IRR=1.49, 95% CI=1.34, 1.59 v. 1.46, 95% CI=1.23, 1.80 respectively).

**Table 3.** Weighted associations between InfLaMeS and health outcomes in the Health and Retirement Study testing sample (N=1799)

|  | **Mortality** |  | **ADLs+IADLs**<br>(number of disabilities) |  | **Multimorbidity** |  |
| --- | --- | --- | --- | --- | --- | --- |
| **Exposure** | **OR**<br>**(95% CI)** | **p-value** | **IRR**<br>**(95% CI)** | **p-value** | **OR**<br>**(95% CI)** | **p-value** |
| InfLaMeS | 2.24<br>(1.80, 2.73) | <0.001 | 1.49<br>(1.34, 1.59) | <0.001 | 1.75<br>(1.52, 2.01) | <0.001 |
| Inflammation latent variable | 1.78<br>(1.52, 2.08) | <0.001 | 1.46<br>(1.23, 1.80) | <0.001 | 1.42<br>(1.25, 1.53) | <0.001 |

Both InfLaMeS and the latent variable are z-scored for comparability. All models adjusted for age, sex, race/ethnicity, smoking status, obesity and alcohol consumption. OR: odds ratio; CI: confidence interval; IRR: incident rate ratio; ADL: activities of daily living; IADL: instrumental activities of daily living

### Replication

In TILDA participants, InfLaMeS was positively correlated with IL-1RA, TNFR-II, CRP and IL-6 (Table 4). Correlation sizes were similar to or slightly smaller than those observed in HRS (e.g., CRP r=0.44; IL-6 r=0.29). No statistically significant correlation was identified with IGF-1 and IL-10. Associations between InfLaMeS and mortality and multimorbidity were replicated in TILDA (Table 5) using logistic regression adjusting for age, sex, smoking status (past, current, never), obesity (not obese, obese, overweight) and alcohol consumption (non-drinker, 5+ drinks, missing). Higher InfLaMeS values were associated with higher odds of 12-year mortality (OR = 1.65, 95% CI: 1.13, 2.43) and higher odds of multimorbidity (OR = 1.66, 95% CI:1.31, 2.11). While the estimate of the association between InfLaMeS and ADLs+IADLs using a negative binomial model due to high overdispersion was elevated, it was not statistically significant (IRR = 1.35, 95% CI: 0.91, 2.02).

**Table 4.**
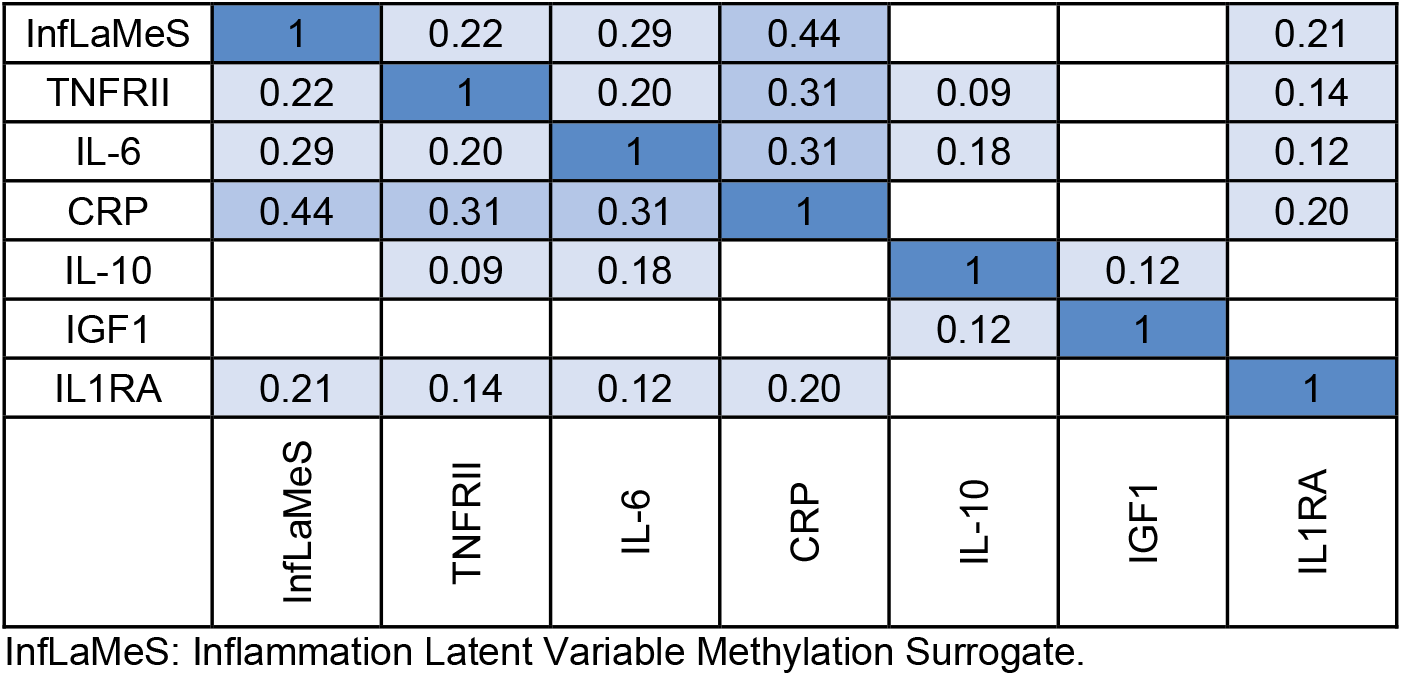
Correlation matrix of InfLaMeS and standardized and log-transformed inflammatory markers in The Irish Longitudinal Study on Ageing, n=488 Insignificant correlations (p>0.05) are blank/white.

**Table 5.** Associations between InfLaMeS and health outcomes of interest in The Irish Longitudinal Study on Ageing (n=488).

|  | **Mortality** |  | **ADLs+IADLs** |  | **Multimorbidity** |  |
| --- | --- | --- | --- | --- | --- | --- |
| **Exposure** | **OR**<br>**(95% CI)** | **p-value** | **IRR**<br>**(95% CI)** | **p-value** | **OR**<br>**(95% CI)** | **p-value** |
| InfLaMeS | 1.65<br>(1.13, 2.43) | 0.01 | 1.35<br>(0.91, 2.02) | 0.12 | 1.66<br>(1.31, 2.11) | <0.0001 |

All models adjusted for age, sex, race/ethnicity, smoking status, obesity and alcohol consumption; OR: odds ratio; CI: confidence interval; IRR: incident rate ratio; ADL: activities of daily living; IADL: instrumental activities of daily living

## DISCUSSION

Chronic inflammation is arguably one of the most important components of aging and age-related disease processes and has been widely used in scientific research to differentiate biological risk for late-life health problems and to evaluate biosocial pathways that contribute to health inequalities. While several scientific studies have incorporated measures of inflammation, there is significant variability in the number and type of inflammatory markers (e.g. CRP, IL-6, IL-10) used across them. Differences in the “operationalization” of inflammation may lead to lack of replication, which can impede scientific progress in understanding the role of inflammation in the development of age-related health. To address this gap, we used DNA methylation data from the HRS to create InfLaMeS, a DNAm surrogate for chronic systemic inflammation, based on a latent inflammation factor established in past research.^10^ InfLaMeS was strongly correlated with measured inflammatory markers and performed similarly to—or slightly better than—the latent systemic inflammation when predicting multimorbidity, disability, and 4-year mortality. Importantly, InfLaMeS was validated using data from TILDA, an independent cohort of older adults. This measure provides a robust alternative to blood-chemistry measures of inflammation that can be used for further scientific insight into understanding the role of inflammation in aging and health.

While epigenetic clocks are among some of the most widely used DNAm surrogates in biomedical and social science research, these measures are broad biomarkers of aging and health – trained to predict several health outcomes years before the onset of clinical symptoms. However, epigenetic clocks may not provide precise insight into how specific health risks or biological processes impact aging and health in later life. To address this gap, recent advancements in Geroscience have led to the creation of multiple surrogates that were trained on specific exposures or health outcomes. For example, DNAm surrogates have been generated to predict short-term cardiovascular event risk^38^, cumulative lead exposure^39^, tobacco smoking^40^, and body mass index (BMI).^41^ These exposure- or outcome-specific DNAm surrogates often perform as well or better than direct measurements of the risk factor for evaluating subsequent health outcomes.^25,26,38^ For example, in the Future of Families and Child Wellbeing Study, a cohort representative of births in US cities, epigenetic BMI at age 9 predicted BMI at age 15 above the measure of BMI at age 9.^41^ In this study, we compare the association between age-related health (e.g. mortality, multimorbidity, and functional limitations) with the latent inflammation marker derived from CRP and cytokine levels and with InfLaMeS. We found that InfLaMeS often performed better than the latent factor of inflammation when predicting subsequent multimorbidity and 4-year mortality. Thus, InfLaMeS may provide a more robust way of examining inflammation-related biological risk for biomedical and social science research.

Additionally, DNAm methylation surrogates are highly portable, which may make them a useful tool for measurement, harmonization, and comparison across studies. Specifically, studies often vary in the number and type of inflammation markers that are collected and used. For example, CRP has been one of the most widely used inflammation markers. However, prior studies have found that CRP may not predict mortality and health as well as a latent construct of inflammation^10^, nor is it always strongly associated with other markers of inflammation.^42^ However, studies may not collect several other inflammation markers due to blood volume, costs, or other research-related limitations. As such, a DNAm surrogate of inflammation provides an alternative to processing additional inflammation markers if DNA methylation data has already been or is planning to be collected, which can also allow for better harmonization and comparison. The replication of InfLaMeS in the TILDA study provides further evidence that supports this potential application. Additionally, a very large number of studies (including many publicly available datasets from resources like the Gene Expression Omnibus) have collected DNAm data but have not collected inflammation markers at all. Tools like InfLaMeS allow researchers to analyze systemic inflammation in these studies.

Lastly, it is also important to point out that DNAm surrogates can provide another useful measure to study the life course origins of health and aging. For example, it has long been theorized that inflammation-related health problems in later life stem from several life course exposures. Studies have often evaluated inflammation at multiple life stages, ranging from childhood to later adulthood. Inflammation will likely look different across these different life course stages– as chronic lifetime inflammation in older adulthood takes years or decades to accumulate.^43^ In contrast, DNAm surrogates for systemic inflammation, such as InfLaMeS, may be more stable than cross-sectional cytokine measurement because DNAm changes may be biologically upstream from blood-chemistry changes and DNAm alterations occur more slowly over time. As such, the DNAm surrogates in later life may provide insight into the potential upregulation of inflammation set in early life and may relate better to chronic exposures. Prior studies have shown that DNAm markers for inflammation created in older adults have been validated in childhood cohorts.^44^ Future research may be able to use InfLaMeS to investigate the early life origins of inflammation-related aging and health outcomes, which may provide further insight into the role of epigenetic change.

While our study is among the first to create a DNAm surrogate of chronic inflammation, which greatly impacts health and aging processes, we acknowledge a few limitations. First, InfLaMeS is based on cross-sectional inflammation information rather than based on longitudinal change. Inflammation is a dynamic process; therefore, future research should consider whether a DNAm surrogate-based change in inflammation may provide additional insight into inflammaging. Second, our health outcomes were based on a limited time frame making our results conservative. Analyses of multimorbidity and ADLs + IADLs were cross-sectional. However, mortality analyses were 4 years and 12 years after blood collection in HRS and TILDA, respectively. Future studies should examine how InfLaMeS is related to health and mortality over a longer follow-up period, as distinct patterns may emerge. Third, our measurement of mortality was based on deaths from all causes. Therefore, the relationship between the InfLaMeS and mortality may be more conservative, as it includes other causes of mortality that may not be as closely linked to inflammation (i.e. suicides, accidents, etc.).

Despite these limitations, our study has strengths. Notably, the HRS is a large nationally representative and diverse sample of US older adults, with many participants that provided DNAm data. Further, we successfully replicated associations between InfLaMeS and health outcomes in an external dataset from a different national and cultural context. InfLaMeS represents a more comprehensive measure of inflammation than prior biomarkers based on individual inflammatory markers.

## CONCLUSION

This study puts forward InfLaMeS, the first DNA methylation surrogate on chronic systemic inflammation, which is a key contributor to health and aging. This surrogate marker provides a next potential step to understanding how systemic inflammation may be impacting health and aging outcomes across various populations. The portability of this measure across studies demonstrates its utility in populations with significantly different social, cultural, and economic histories. As such, it can be used to better understand the causes and consequences of inflammation in populations across the world.

## Data Availability

HRS data are available on the HRS website [https://hrs.isr.umich.edu/] and in the HRS NIAGADS repository [https://dss.niagads.org/datasets/ng00153/]. TILDA data are available from TILDA [https://tilda.tcd.ie/data/accessing-data/].

https://dss.niagads.org/datasets/ng00153/

https://hrs.isr.umich.edu/data-products

## FUNDING

This work was supported by U01AG073289, R01AG068937, R01AG060110, and R01AG071071. The Health and Retirement Study is sponsored by the National Institute on Aging (U01AG009740) and conducted by the University of Michigan. SFI US-Ireland Research and Development Partnership (Grant Number 19/US/3615) (BH, CMcC, RAK). TILDA is supported by the Irish Government, the Atlantic Philanthropies, and the Health Research Board.

## Notes

### Competing Interest Statement

The authors have declared no competing interest.

